# Evaluating Patient Experience in a Rural-Serving Nephrology Clinic: Psychometric assessment of the consideRATE questions and assessment of care experience

**DOI:** 10.1101/2025.04.27.25326517

**Authors:** Joseph P. Nano, Anji Zhu, Rhea Sachdeva, Ann M. O’Hare, Annika Milliman, Clay A. Block, Glyn Elwyn, JoAnna K. Leyenaar, Lorraine Gerraughty, Meredith A. MacMartin, Nicholas M. Fuller, Steven L. Bernstein, Vanessa L. Junkins, Catherine H. Saunders

**Affiliations:** Dartmouth Geisel School of Medicine, Hanover, NH, USA; Dartmouth Hitchcock Medical Center, Lebanon, NH, USA; University of Washington, Seattle, WA, USA

**Keywords:** **conside**RATE, Patient experiences, Primary care, Rural health, Rurality, Psychometric

## Abstract

**Background:** The illness experiences of people with kidney disease are not systematically ascertained as part of clinical care. We conducted the first real-world psychometric evaluation of the **conside**RATE questions among patients with kidney disease and assessed healthcare experiences in a rural hospital setting.

**Methods:** We conducted a cross-sectional survey with a sample of patients with kidney disease and their care partners. Our survey consisted of demographic questions, **conside**RATE (8-items) to measure healthcare experience, and CANHELP-Lite (21-items) as a reference for **conside**RATE psychometric assessment. We scored **conside**RATE as continuous (range: 1-4) and top-box (score=4). We conducted internal reliability (Cronbach’s alpha) and validity (convergent and discriminant using Pearson’s correlation) tests. We compared mean **conside**RATE scores between paired subgroups using t-tests. We examined the relationship between rurality and healthcare experience using a multivariable linear regression model and rural-urban commuting area codes.

**Results:** We recruited 163 participants, including 52 who lived in rural areas. The mean **conside**RATE score was 3.65 (SD=0.42), and the top-box score was 35% (n=59). Internal consistency for **conside**RATE was 0.86. We demonstrated convergent validity (continuous: r=0.5, p<0.001 and top-box: r=0.3, p<0.001) and discriminant validity (continuous: r=0.1, p>0.05 and top-box: r=0.1, p>0.05). Subgroup analyses revealed no significant differences in overall **conside**RATE scores by rurality status, state of residence, travel time, and gender. After adjustment for demographics, lower continuous mean **conside**RATE scores were significantly associated with longer travel time to the clinic (p<0.01), religious affiliation (p<0.05), and age over 55 years (p<0.01). We found no significant association between total **conside**RATE score and rurality.

**Conclusion:** We found validity in the first real-world psychometric assessment of **conside**RATE among people with kidney disease and found associations between healthcare experience and various demographic variables.

## Background

With population growth and aging, patients with serious illnesses, such as cancer, heart disease, and chronic kidney disease (CKD), have unmet needs that affect the quality of their care and their experiences of illness and care.^1,2^ Approximately 35.5 million adults in the United States (US) (more than 1 in 7 US adults) and over 700 million people globally (10% of the world population) are affected by CKD.^3,4^ CKD is a costly, resource intensive disease to manage.^4^ In 2019, treatment for Medicare beneficiaries with CKD cost $87.2 billion even without including the costs for dialysis or transplant.^4^ CKD is often a silent disease until advanced stages when patients experience a variety of symptoms that affect their health-related quality of life.^5,6^ Furthermore, patients with kidney disease, along with their care partners, may face many barriers to receiving healthcare services particularly in rural areas, such as frequent follow-up appointments, long-distance travel, and the financial burden associated with treatments and related expenses.^7,8^ This population is often underrepresented in research, with most studies focusing on urban-rural settings outside of the United States.^9,10^ As a result, less is known about the healthcare experiences and needs of rural patients with CKD.

Patient-reported experience measures (PREMs) provide a means to capture objective accounts of patients’ experiences and their satisfaction with care.^11–13^ PREMs are increasingly utilized to monitor experiences in various populations because they provide valuable insights into the patient’s perspective on their health, treatment, and overall well-being.^11,12^ This is important because better patient experience has been linked to improved clinical outcomes, including greater adherence to treatment, reduced hospital readmissions, and higher quality of care.^14^ Furthermore, increased patient engagement can empower patients to self manage conditions and positively impact clinical and financial outcomes by improving clinical care benchmarks and reducing costly resource utilization. The **conside**RATE questions, a measure of serious illness care experience, provide meaningful measurement of patient-reported experiences and can help to identify opportunities to improve the experience of patients and care partners, across the illness trajectory.^15^ Unlike other measures (e.g., the CANHELP Lite measure), the **conside**RATE questions are a novel, patient-centered tool with a short completion time.^15–19^ This tool was validated in both a simulated online setting and among patients with cancer.^16,19^ Furthermore, the **conside**RATE questions have the potential to enhance routine care for patients with kidney disease through providing valuable real-time feedback to care teams, helping to inform and improve care delivery.^15,19,20^

There are significant gaps in the literature regarding the association between rurality and patient experience. For example, while several studies have investigated patient experiences in rural and urban areas, the findings have been inconsistent.^21–24^ Furthermore, the real-world psychometric properties of the **conside**RATE questions in non-cancer clinical settings and populations—such as patients with kidney disease—remain unknown. To address these gaps, we conducted a study to examine the association between rurality and patient experience in a rural-serving clinic in the US. This is also the first real-world assessment of the psychometric properties of **conside**RATE in a population with kidney disease. We hypothesized that healthcare experience will vary across rurality status due to geographic isolation, limited financial resources, and lower socioeconomic status.

## Materials and Methods

### Design

We conducted a single-time point cross-sectional survey among a convenience sample of patients and their care partners attending a single nephrology clinic at Dartmouth-Hitchcock Medical Center (DHMC), located in Lebanon, NH. DHMC is a tertiary care center located in rural Northern New England, and is also New Hampshire’s only academic medical center. The Dartmouth Health Institutional Review Board (IRB) approved this study in May 2023 [IRB ID: 02000560]. We reported results using the Checklist for Reporting of Survey Studies (CROSS).

### Participants

We included patients or care partners visiting the nephrology clinic at Dartmouth-Hitchcock Medical Center in Lebanon, New Hampshire, which serves a largely rural population. Participants were required to be older than 18, be able to read English, and provide written informed consent to participate in the study. Some participant responses were excluded from the final analysis due to interruptions in completing the measures in the waiting room of the nephrology clinic.

### Survey validation

Using Qualtrics, an online survey platform, we designed a 15-minute survey consisting of 51 questions. The survey included a demographic survey, the **conside**RATE questions, and the CANHELP Lite questionnaire.^18^ We designed the survey and analytic plan based on a previously conducted online validation study, which assessed reliability and (convergent, discriminative, and divergent) validity.^16^ Before distributing the survey to participants, we piloted the survey with members of our team to ensure it was functioning well and to identify and address any technical, design or clarity issues.

### Survey elements

#### Demographic Survey

Our survey consisted of demographic questions (checkbox), such as identity (patient or accompanying care partner), age, race, gender, ethnicity, disease stage (eGFR), rurality, and list of comorbidities (self-reported by respondents) (**Table 1**).

**Table 1.**
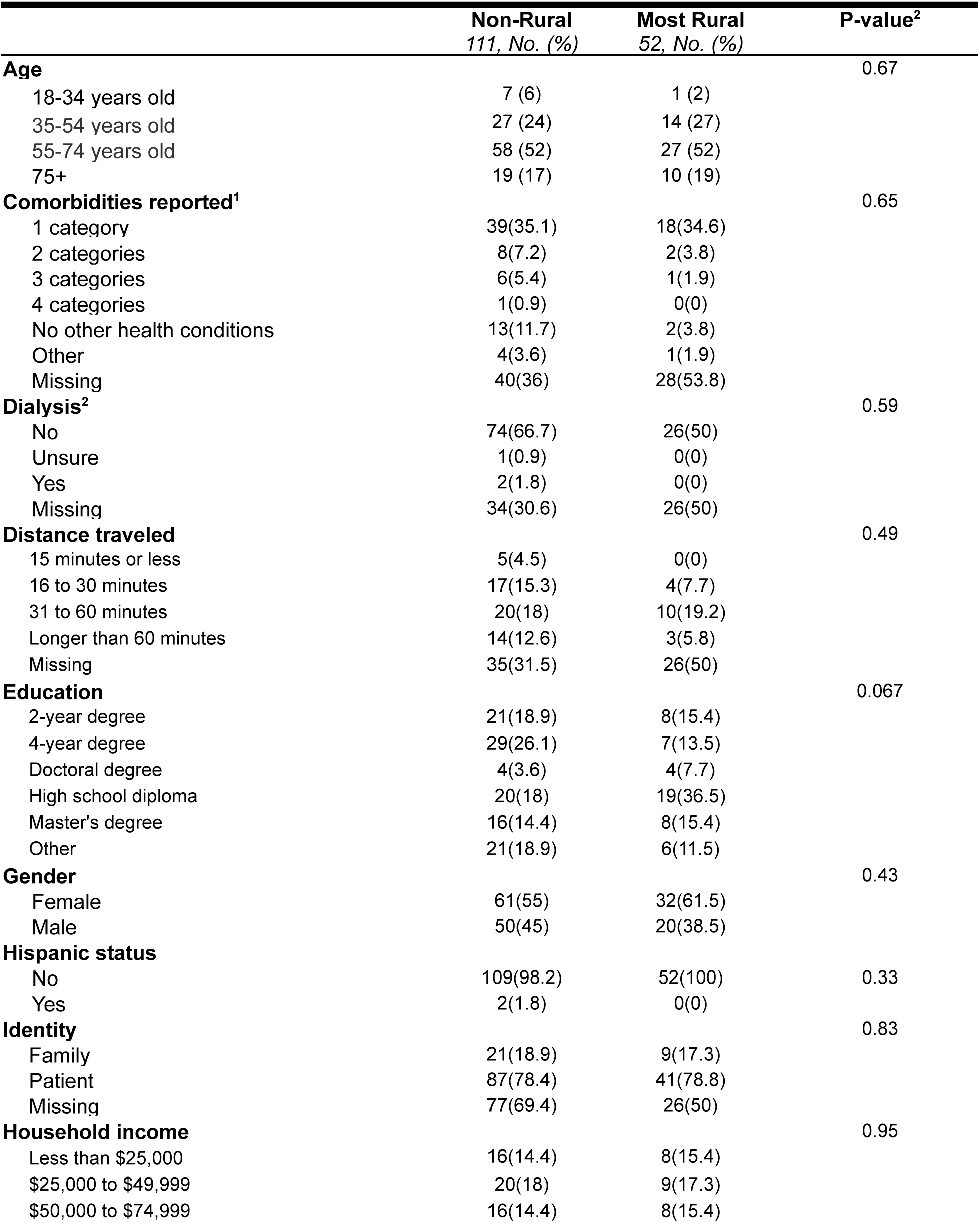
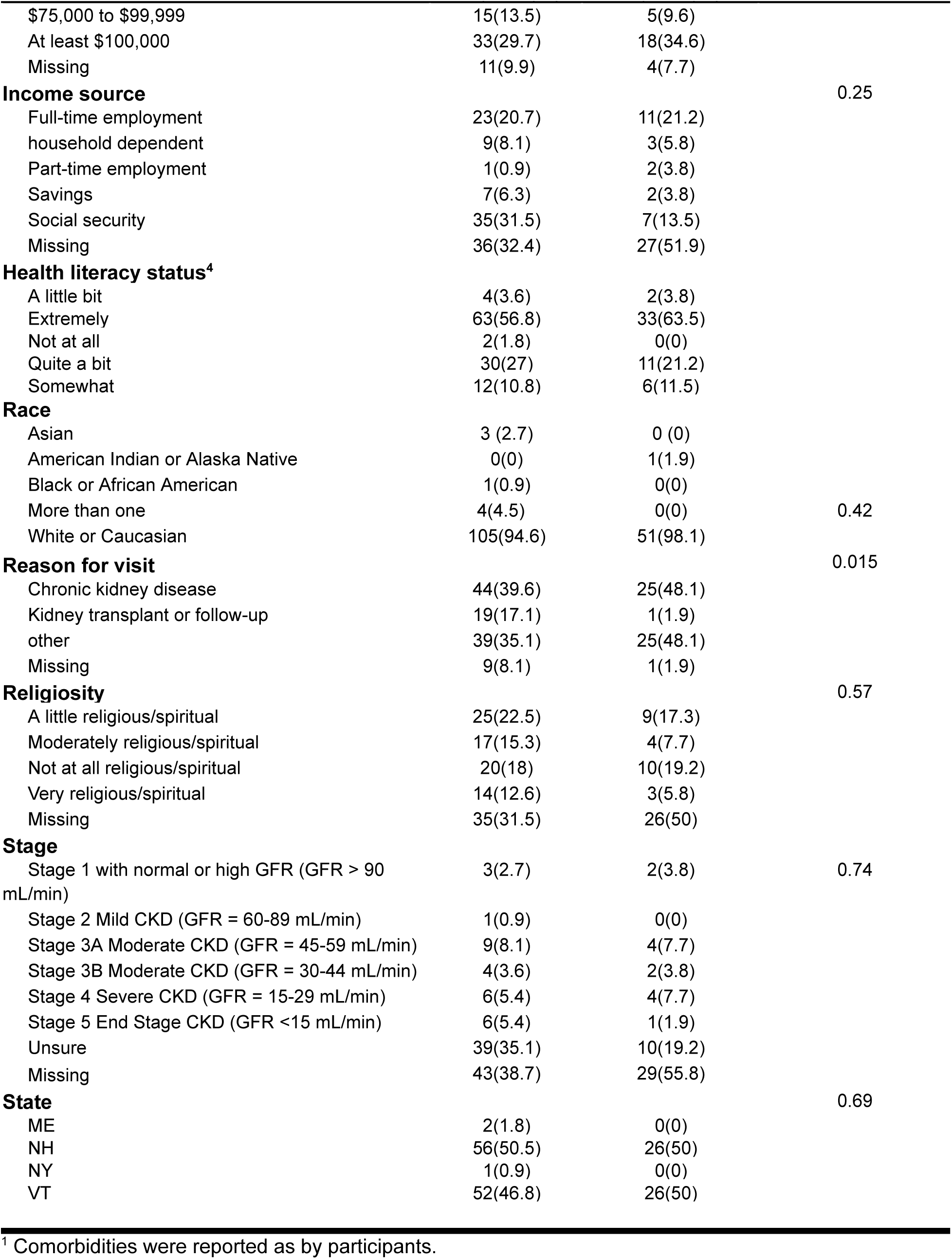
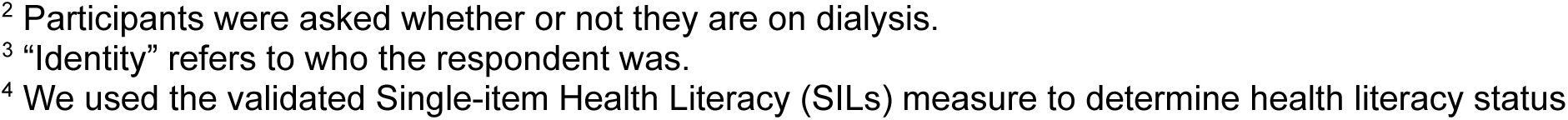
Psychometric tests using overall instrument scoring^1^.

#### The consideRATE questions

The **conside**RATE is an 8-item self-reported measure of serious illness care experiences for patients and their care partners that can be administered in various clinical settings, such as outpatient, inpatient, and home care.^15^ The first seven questions are scored on a four-point Likert-like scale (1 = very bad, 2 = bad, good = 3, and 4 = very good) and the last question is an open-ended question. We excluded an optional ninth-item regarding desired anonymity as it was inappropriate in a research versus quality improvement context.

#### CANHELP Lite

CANHELP Lite (Canadian Health Care Evaluation Project) is a 21-item patient-facing and validated measure of serious illness care experience that is scored on a five-point Likert scale (1 = Not at all, 2 = Not very, 3 = Somewhat, 4 = Very, and 5 = Completely).^18^ We selected CANHELP Lite for consistency as we used this instrument in two previous consideRATE validation studies for cancer patients and older adults. We conducted the previous two validation studies of consideRATE, in older adults, and cancer patients, using this instrument.^16,19^ We used CANHELP Lite as a continuous measure for the psychometric assessment of **conside**RATE given the similarity in response options.

### Procedures

Between February 2024 and October 2024, we recruited patients with kidney disease and care partners or family members who accompanied patients. Using convenience sampling, we randomly approached participants after they checked in at the nephrology clinic main desk. Our recruitment team explained the purpose of the study, answered questions, and obtained verbal and written consent before providing the survey.

### Statistical analysis

We used Microsoft Excel to perform data cleaning and conducted statistical analyses using R-Studio statistical software (version 1.3.959, 2009–2020 R-Studio, PBC). In the first phase of analysis, we used simple descriptive statistics to summarize the demographics of the sample. In the second phase of analysis, we validated our **conside**RATE measure in the kidney disease population by testing reliability, convergent validity, and discriminant validity. We defined weak correlation as less than r=0.3, moderate between r=0.3 to r=0.7, and strong correlation as at least r=0.7.^25^ Third, we fitted a multivariable regression model to identify predictors of healthcare experience using **conside**RATE for healthcare experience, Rural-Urban Commuting Area (RUCA) codes, and other demographic variables. We assigned RUCA codes to participants based on their residential ZIP codes, allowing us to assess the impact of rurality on healthcare experiences. We classified rurality based on the Rural Urban Commuting Area four-tier classification schema: Metropolitan area, Micropolitan area, Small town area, and Rural area.^26^ The model was adjusted for key demographic characteristics such as age, gender, race/ethnicity, education, and travel time to the clinic. We set the significance level at α = 0.05 and did not adjust for multiple comparisons, as this was an exploratory analysis. Fourth, we conducted a content analysis of participants’ responses to the **conside**RATE open-ended question.

#### Psychometric assessment of consideRATE

##### I. Overall Instrument Scoring

We used two scoring approaches to test the convergent validity of the **conside**RATE questions that were performed in previous studies: continuous scoring and top-box scoring. For the continuous scoring approach, we used the original **conside**RATE four-point and CANHELP Lite five-point scales and calculated mean scores for both measures across all items (seven-items for **conside**RATE and 21-items for CANHELP Lite). Then we compared the mean score of **conside**RATE with the mean score of CANHELP Lite for each participant.

##### II. Top-box Scoring

For top-box scoring, we dichotomized the **conside**RATE overall mean score (1 = “very good”; 0 = “good” or “bad” or “very bad”). We compared the top-box dichotomized **conside**RATE overall mean score with the original continuous CANHELP Lite scoring using a point-biserial correlation test.

##### III. Item-by-item Scoring

To perform item-by-item convergent validity tests, we used a similar approach that we performed in a previous validation study, in which we conducted Pearson’s correlation tests on the matched individual items using the **conside**RATE scale to items on the CANHELP Lite scale.^19^ For discriminant validity tests, we identified a measure that is conceptually distinct from the serious illness experience measured in **conside**RATE, the validated Single-item Health Literacy (SILs) measure.^27–30^ This instrument asks “How confident are you in filling out medical forms by yourself?” and offers the following response options: “Extremely”, “Quite a bit”, “Somewhat”, “A little bit”, “Not at all.” We selected SILs because it does not directly address **conside**RATE constructs and does not overlap conceptually with any of the seven **conside**RATE questions.

We conducted Pearson’s correlation tests between the **conside**RATE scale and SILs measure. We expected moderate to strong significant correlations for convergent validity and weak correlations to no correlation for discriminant validity.^25^

##### IV. Reliability

We conducted a reliability test on the **conside**RATE questions using the Cronbach alpha reliability test. We considered “good” internal consistency for Cronbach’s alpha score to be 0.7 or higher.^31^

#### Subgroup analysis

We used t-tests to compare mean **conside**RATE scores between four paired subgroups based on state of residence (New Hampshire vs. Vermont), reason for visit (CKD vs transplant patient or other), participant’s identity (patient vs. care partner), travel time (less than 60 minutes vs at least 60 minutes), and Rurality status (rural area vs non-rural residing area). We selected these characteristics for subgroup analysis because they were the most commonly reported across participants. Characteristics with very few responses (e.g., for state, we excluded Maine = 2 and New York = 1) were excluded from subgroup comparisons to avoid unreliable estimates.

#### Multivariable Regression Model

We investigated predictors of healthcare experience by exploring factors associated with participants’ healthcare experience as measured by the **conside**RATE tool using two approaches: (1) using the total **conside**RATE score as the dependent variable and (2) using the **conside**RATE score per item as the dependent variable (e.g., “Physical problems” item, “Emotional problems” item, “What matters most” item, “Surroundings” item). First, we used a linear regression model for the continuous total **conside**RATE score. Second, given that the **conside**RATE measure consists of seven items, each capturing a distinct aspect of the patient healthcare experience, we conducted a series of linear regression models across each item to investigate associations between each item with various demographic and clinical characteristics. We selected covariates by known associations between healthcare experience and rurality based on previous literature conducted outside of the United States.^21,22,24^ We investigated whether “distance traveled” is a confounding variable for the association between healthcare experience and independent variables before including it as a covariate.^32,33^ We checked for multicollinearity of the model using Variance Inflation Factor (VIF). We found the adjusted generalized standard error inflation factor (aGSIF) across variables to range between 1.06 and 1.16, which shows that the model’s coefficient estimates are stable and reliable.

#### ConsideRATE: Open-ended question

We conducted a qualitative content analysis to examine participants’ responses to the **conside**RATE open-ended question in our survey. Two researchers (AZ and RS) read responses and captured key concepts independently. Using an inductive approach, both researchers applied categorical codes to extract meaning from the text. We synthesized and refined codes into broader categories. We reviewed categories to ensure they were consistent with the coded data and accurately represented the overall dataset.

Differences in interpretation were resolved by discussion.

## Results

### Descriptive Statistics

We approached 312 potential participants and 256 participants agreed to participate. A total of 163 (52%) of those approached had time to provide informed consent and complete the survey (**conside**RATE mean score = 3.65, SD = 0.42). Of these, 111 (68%) reported that they reside in non-rural areas (mean score = 3.68, SD = 0.40) and 52 (32%) in rural areas (mean score = 3.61, SD = 0.41) (**Table 1**) (**Supplementary 1**). Approximately 45% (n = 69) reported visiting nephrology clinics for a “chronic kidney disease” appointment, 13% (n = 20) for “transplant or follow up”, and 42% (n = 64) “other” (e.g., first appointment, referral). Most study participants were patients (n=128, 79%) and the remainder were family members or care partners (n=30, 18%).

Approximately half of the participants were female (n=93, 57%) and most identified as white or Caucasian (n=156, 96%) (**Table 1**) and non-Hispanic (n=161, 99%). Over half of the participants (n=85, 52%) were aged between 55–74 years, (n=29, 18%) 75+ years, with a minority (n=8, 5%) aged 18–34 years old. Among participants who reported residing in rural areas, approximately 20% reported traveling 31 to 60 minutes for care, while 6% traveled longer than 60 minutes (**Table 1**).

### Psychometric assessment of consideRATE

#### Overall instrument: Convergent validity

Among all participants, we found a statistically significant moderate correlation (r=0.5; *p*<0.001) between the overall continuous scores of the **conside**RATE questions and CANHELP Lite, indicating adequate convergent validity (**Table 2**). For top-box scoring among all participants, we found a statistically significant moderate correlation (r^pb^=0.3; *p*<0.001) between the overall top-box scoring of the **conside**RATE questions and the CANHELP Lite continuous scores, indicating adequate convergent validity (**Table 2**). In subgroup analyses, we found moderate to strong correlations between **conside**RATE scores and CANHELP Lite scores in patients, care partners, those with low-income, those with low educational attainment, those residing in rural areas, those visiting for CKD appointments, and those identified as transplant patients. These correlations indicate strong convergent validity for continuously-scored **conside**RATE for these select groups (r range: 0.6 to 0.8, p<0.001) and top-box (r range: 0.4 to 0.6, p<0.01) (**Table 2**) scores.

**Table 2.** Psychometric tests using overall instrument scoring^1^.

| Convergent validity | All participants<br>r (p-value) | Patients only<br>r (p-value) | Care Partners only<br>r (p-value) | Low-income<br>r (p-value) | Low educational attainment<br>r (p-value) | Rural area<br>r (p-value) | Chronic kidney disease<br>r (p-value) |
| --- | --- | --- | --- | --- | --- | --- | --- |
| Mean **considerate** questions scores vs. mean CANHELP Lite score | 0.5***<br>(<0.001) | 0.6***<br>(<0.001) | 0.7***<br>(<0.001) | 0.8***<br>(<0.001) | 0.7***<br>(<0.001) | 0.7***<br>(<0.001) | 0.6***<br>(<0.001) |
| Top-box dichotomous **considerate** questions scores vs. mean CANHELP Lite score | 0.3***<br>(<0.001) | 0.4***<br>(<0.001) | 0.3<br>(0.087) | 0.5***<br>(0.004) | 0.4*<br>(0.036) | 0.6***<br>(<0.001) | 0.4***<br>(0.001) |
| Reliability | Mean (SD) | Standard deviation | Scale Variance if Item Deleted | Corrected item-Total correlation | Squared Multiple Correlation | Cronbach's Alpha if Item Deleted |  |
| **Overall Cronbach alpha = 0.89** |  |  |  |  |  |  |  |
| Physical item | 3.67 | 0.53 | 6.70 | 0.71 | 0.53 | 0.87 |  |
| Surrounding item | 3.71 | 0.63 | 6.33 | 0.70 | 0.53 | 0.87 |  |
| Feelings item | 3.75 | 0.48 | 6.74 | 0.79 | 0.69 | 0.86 |  |
| Matters item | 3.73 | 0.52 | 6.55 | 0.78 | 0.69 | 0.86 |  |
| Plan item | 3.66 | 0.51 | 6.90 | 0.65 | 0.48 | 0.88 |  |
| Affairs item | 3.62 | 0.52 | 7.15 | 0.53 | 0.32 | 0.89 |  |
| Expect item | 3.62 | 0.64 | 6.33 | 0.68 | 0.49 | 0.88 |  |

#### Item-by-item scoring: Convergent and Discriminant Validity

Among all participants, we found moderate statistically significant correlations ranging from r=0.3 (*p*<0.001) to 0.5 (*p*<0.001) across all individual matched **conside**RATE continuous scoring and CANHELP Lite items, indicating moderate convergent validity (**Table 3**). For top-box scoring among all participants, we found moderate statistically significant correlations ranging from r^pb^=0.3 (*p*<0.001) to r^pb^=0.5 (*p*<0.001) across all individual questions, indicating convergent validity for all items (**Table 3**). For discriminant validity, we found no significant correlation between **conside**RATE healthcare items and the demographic literacy item for continuous **conside**RATE (r range: r=0.07 to r=0.07, p>0.05) and top-box **conside**RATE (r range: r=0.001 to r=0.147, p>0.05) (**Table 3**). We found a significant correlation with low Pearson’s correlation coefficient between **conside**RATE and the demographic literacy question across a few items for continuous **conside**RATE (r=0.2, p<0.05) and top-box **conside**RATE (r range: r=0.1 to 0.2, p<0.05) (**Table 3**).

**Table 3.** Matching consideRATE questions with CANHELP Lite and demographic questions for convergent and discriminant validity^1^.

| Convergent Validity |  | ConsiderATE |  |  |
| --- | --- | --- | --- | --- |
| Construct Summary<br>Care of | ConsiderATE questions | CANHELP Lite questions | Continuous<br>r (p-value) | Top-box<br>r (p-value) |
| Physical problems | Q1: "How would you rate our attention to your physical problems? Things like pain, dry mouth or trouble breathing" | Q1: "How satisfied are you that physical symptoms you had (for example: pain, shortness of breath, nausea) were adequately assessed and controlled?" | 0.5***<br>(<0.001) | 0.5***<br>(<0.001) |
| Emotional problems | Q2: "How would you rate our attention to your feelings? Things like feeling sad, worried, or like a burden" | Q2: "How satisfied are you that emotional problems you had (for example: depression, anxiety) were adequately assessed and controlled?" | 0.3***<br>(<0.001) | 0.3***<br>(<0.001) |
| Surrounding problems | Q3: "How would you rate our attention to your surroundings? Things like noise, light, or warmth" | Q3: "How satisfied are you with the environment or the surroundings in which you were cared for?" | 0.4***<br>(<0.001) | 0.4***<br>(<0.001) |
| What matters most | Q4: "How would you rate our respect for what matters to you? Things like values, preferences about care, or important activities" | Q4: "How satisfied are you that you received consistent information about your condition from all doctors and nurses looking after you?" | 0.3***<br>(<0.001) | 0.4***<br>(<0.001) |
| Plans | Q5: "How would you rate our communication about your plans? Things like medicines, procedures or place of care" | Q5: "How satisfied are you with discussions with your doctor(s) about where you would be cared for (in hospital, at home, or elsewhere) if your condition worsened?" | 0.3***<br>(<0.001) | 0.3***<br>(<0.001) |
| Affairs | Q6: "How would you rate our attention to your affairs? Things like a will, finances, or advance directives for care" | Q6: "How satisfied are you that you were able to manage the financial costs associated with your illness?" | 0.3***<br>(<0.001) | 0.3***<br>(<0.001) |
| What to expect | Q7: "How would you rate our communication about what you can expect? Things like illness getting worse or time left to live" | Q7: "How satisfied are you with discussions with your doctor(s) about the use of life-sustaining technologies (for example: CPR or cardiopulmonary resuscitation, breathing machines, dialysis)?" | 0.3**<br>(0.005) | 0.3***<br>($<0.001$) |
| **Discriminant Validity** |  |  |  |  |
| Physical problems | Q1: "How would you rate our attention to your physical problems? Things like pain, dry mouth or trouble breathing" | Q1: "How confident are you in filling out medical forms by yourself?" | 0.2*<br>(0.013) | 0.1<br>(0.059) |
| Emotional problems | Q2: "How would you rate our attention to your feelings? Things like feeling sad, worried, or like a burden" | Q1: "How confident are you in filling out medical forms by yourself?" | 0.2*<br>(0.027) | 0.2*<br>(0.043) |
| Surrounding problems | Q3: "How would you rate our attention to your surroundings? Things like noise, light, or warmth" | Q1: "How confident are you in filling out medical forms by yourself?" | 0.1<br>(0.409) | 0.001<br>(0.991) |
| What matters most | Q4: "How would you rate our respect for what matters to you? Things like values, preferences about care, or important activities" | Q1: "How confident are you in filling out medical forms by yourself?" | 0.2***<br>(0.006) | 0.2*<br>(0.023) |
| Plans | Q5: "How would you rate our communication about your plans? Things like medicines, procedures or place of care" | Q1: "How confident are you in filling out medical forms by yourself?" | 0.2*<br>(0.006) | 0.2*<br>(0.014) |
| Affairs | Q6: "How would you rate our attention to your affairs? Things like a will, finances, or advance directives for care" | Q1: "How confident are you in filling out medical forms by yourself?" | 0.1<br>(0.309) | 0.02<br>(0.836) |
| What to expect | Q7: "How would you rate our communication about what you can expect? Things like illness getting worse or time left to live" | Q1: "How confident are you in filling out medical forms by yourself?" | 0.2*<br>(0.014) | 0.2***<br>(0.009) |

<sup>1</sup> To compute correlation coefficients we used Pearson correlation tests for continuous scoring and point-biserial correlation tests for top-box scoring

#### Reliability

The **conside**RATE questions demonstrated reliability with an internal consistency of 0.9 (**Table 2**).

### Subgroup analysis

We found no significant difference in overall **conside**RATE scores between the following subgroups: participants who resided in rural areas and non-rural areas (p=0.362), participants from New Hampshire and Vermont (p = 0.269), patient and care partners (p = 0.307), male and female groups (p = 0.254), CKD stage and transplant or follow-up group (p = 0.199), and those who traveled less than 60 minutes and those who traveled at least 60 minutes (p = 0.080) (**Table 6**).

### Multivariable Regression Model

#### Total **conside**RATE score

The multivariable regression analysis revealed that residing in “rural areas” was not significantly associated with total **conside**RATE score compared to those residing in “metropolitan areas.” The crude and adjusted coefficients were -0.04 (p-value = 0.758) and -0.07 (p-value = 0.153), respectively. We found a positive association between the total **conside**RATE score and participants who were “55-74 years old” (crude coefficient: 0.43, p<0.001 and adjusted coefficient: 0.49, p<0.05) and “at least 75 years old”, compared to those aged “18-34 years” (**Table 4**). Also, we found a positive association between the total **conside**RATE score and those who “traveled 31-60 minutes” and “longer than 60 minutes” for their appointment (crude coefficient: 0.27, p<0.05 and adjusted coefficient: 0.27, p<0.05), compared to those who traveled “less than 30 minutes” (**Table 4**). We found a weak negative association between the total **conside**RATE score and those who reported being “moderately religious” (crude coefficient: -0.23, p>0.05 and adjusted coefficient: -0.27, p<0.05) compared to those who reported being “slightly religious” (**Table 4**).

**Table 4.**
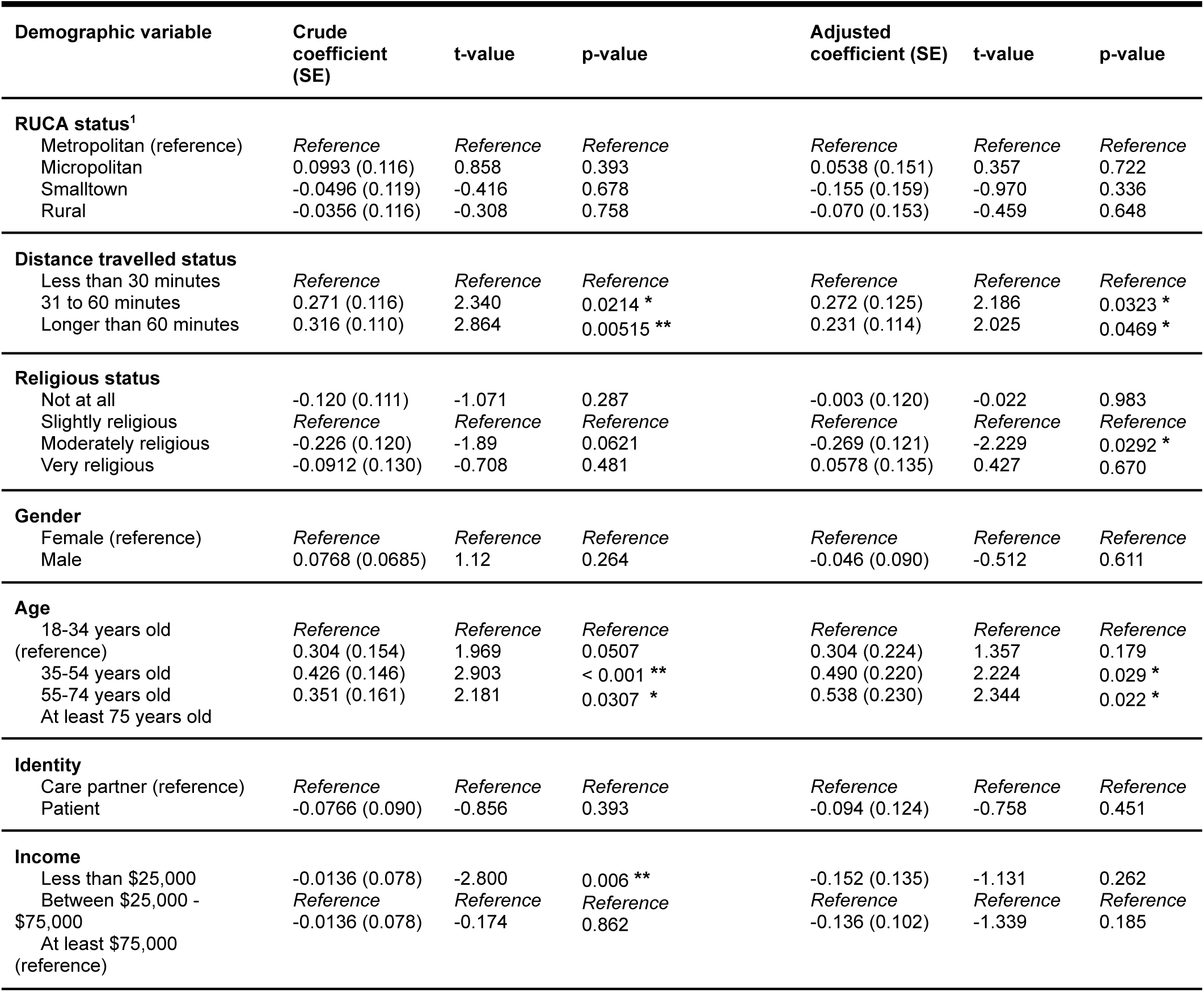
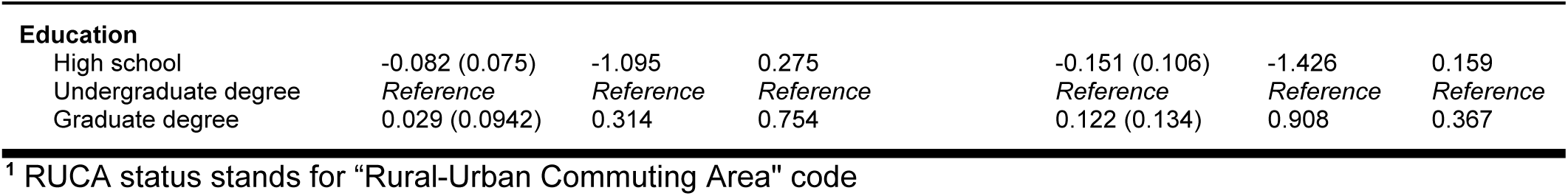
Multivariable linear regression analysis: Association between overall mean consideRATE and demographic variables.

| Demographic variable | Crude coefficient (SE) | t-value | p-value | Adjusted coefficient (SE) | t-value | p-value |
| --- | --- | --- | --- | --- | --- | --- |
| **RUCA status<sup>1</sup>** |  |  |  |  |  |  |
| Metropolitan (reference) | Reference | Reference | Reference | Reference | Reference | Reference |
| Micropolitan | 0.0993 (0.116) | 0.858 | 0.393 | 0.0538 (0.151) | 0.357 | 0.722 |
| Smalltown | -0.0496 (0.119) | -0.416 | 0.678 | -0.155 (0.159) | -0.970 | 0.336 |
| Rural | -0.0356 (0.116) | -0.308 | 0.758 | -0.070 (0.153) | -0.459 | 0.648 |
| **Distance travelled status** |  |  |  |  |  |  |
| Less than 30 minutes | Reference | Reference | Reference | Reference | Reference | Reference |
| 31 to 60 minutes | 0.271 (0.116) | 2.340 | 0.0214 * | 0.272 (0.125) | 2.186 | 0.0323 * |
| Longer than 60 minutes | 0.316 (0.110) | 2.864 | 0.00515 ** | 0.231 (0.114) | 2.025 | 0.0469 * |
| **Religious status** |  |  |  |  |  |  |
| Not at all | -0.120 (0.111) | -1.071 | 0.287 | -0.003 (0.120) | -0.022 | 0.983 |
| Slightly religious | Reference | Reference | Reference | Reference | Reference | Reference |
| Moderately religious | -0.226 (0.120) | -1.89 | 0.0621 | -0.269 (0.121) | -2.229 | 0.0292 * |
| Very religious | -0.0912 (0.130) | -0.708 | 0.481 | 0.0578 (0.135) | 0.427 | 0.670 |
| **Gender** |  |  |  |  |  |  |
| Female (reference) | Reference | Reference | Reference | Reference | Reference | Reference |
| Male | 0.0768 (0.0685) | 1.12 | 0.264 | -0.046 (0.090) | -0.512 | 0.611 |
| **Age** |  |  |  |  |  |  |
| 18-34 years old (reference) | Reference | Reference | Reference | Reference | Reference | Reference |
| 35-54 years old | 0.304 (0.154) | 1.969 | 0.0507 | 0.304 (0.224) | 1.357 | 0.179 |
| 55-74 years old | 0.426 (0.146) | 2.903 | < 0.001 ** | 0.490 (0.220) | 2.224 | 0.029 * |
| At least 75 years old | 0.351 (0.161) | 2.181 | 0.0307 * | 0.538 (0.230) | 2.344 | 0.022 * |
| **Identity** |  |  |  |  |  |  |
| Care partner (reference) | Reference | Reference | Reference | Reference | Reference | Reference |
| Patient | -0.0766 (0.090) | -0.856 | 0.393 | -0.094 (0.124) | -0.758 | 0.451 |
| **Income** |  |  |  |  |  |  |
| Less than \$25,000 | -0.0136 (0.078) | -2.800 | 0.006 ** | -0.152 (0.135) | -1.131 | 0.262 |
| Between \$25,000 - \$75,000 | Reference | Reference | Reference | Reference | Reference | Reference |
| At least \$75,000 (reference) | -0.0136 (0.078) | -0.174 | 0.862 | -0.136 (0.102) | -1.339 | 0.185 |
| **Education** |  |  |  |  |  |  |
| High school | -0.082 (0.075) | -1.095 | 0.275 | -0.151 (0.106) | -1.426 | 0.159 |
| Undergraduate degree | *Reference* | *Reference* | *Reference* | *Reference* | *Reference* | *Reference* |
| Graduate degree | 0.029 (0.0942) | 0.314 | 0.754 | 0.122 (0.134) | 0.908 | 0.367 |

<sup>1</sup> RUCA status stands for "Rural-Urban Commuting Area" code

#### **Conside**RATE score per item

The multivariable linear regression models across each of the seven **conside**RATE items revealed significant associations between three total **conside**RATE score items (the **conside**RATE “Plans” item, the “Surroundings” item, and the “What matters most” item) and various demographic and clinical characteristics (**Supplementary 2**). First, we found a weak negative association between the total **conside**RATE score for the “plans” item and those who have a “high school diploma or less” (crude coefficient: -0.17, p>0.05 and adjusted coefficient: -0.29, p<0.05), compared to those who have an undergraduate degree (**Supplementary 2**). Second, we found a weak negative association between the total **conside**RATE score for the “Surroundings” item for patients (crude coefficient: -0.164, p>0.05 and adjusted coefficient: -0.33, p<0.05), compared to “care partners” (**Supplementary 2**). Third, we also found a weak negative association between the total **conside**RATE score for the “What matters most” item for patients (crude coefficient: -0.164, p>0.05 and adjusted coefficient: -0.33, p<0.05), compared to “care partners” (**Supplementary 2**).

### consideRATE Open-ended Question

A total of 23 patients and care partners provided free-text responses to the **conside**RATE open-ended questions, which underwent screening (by AZ and RS) to identify common patterns and categories, which consisted of positive and negative experiences among patients and care partners. Among the responses, 8 were negative, 10 were positive, and 5 were deemed irrelevant because the participants answered “no” or their responses indicated it was their first visit and they had insufficient information to answer the questions (**Table 5**). Any discrepancies in screening were resolved through discussion among the research team, including senior author CHS. Using a qualitative content analysis approach, AZ and RS conducted an open coding approach of the responses, which were then categorized into five categories: Caring, Scheduling, Personalization, Quality, and Collaboration (**Table 5**).

**Table 5.**
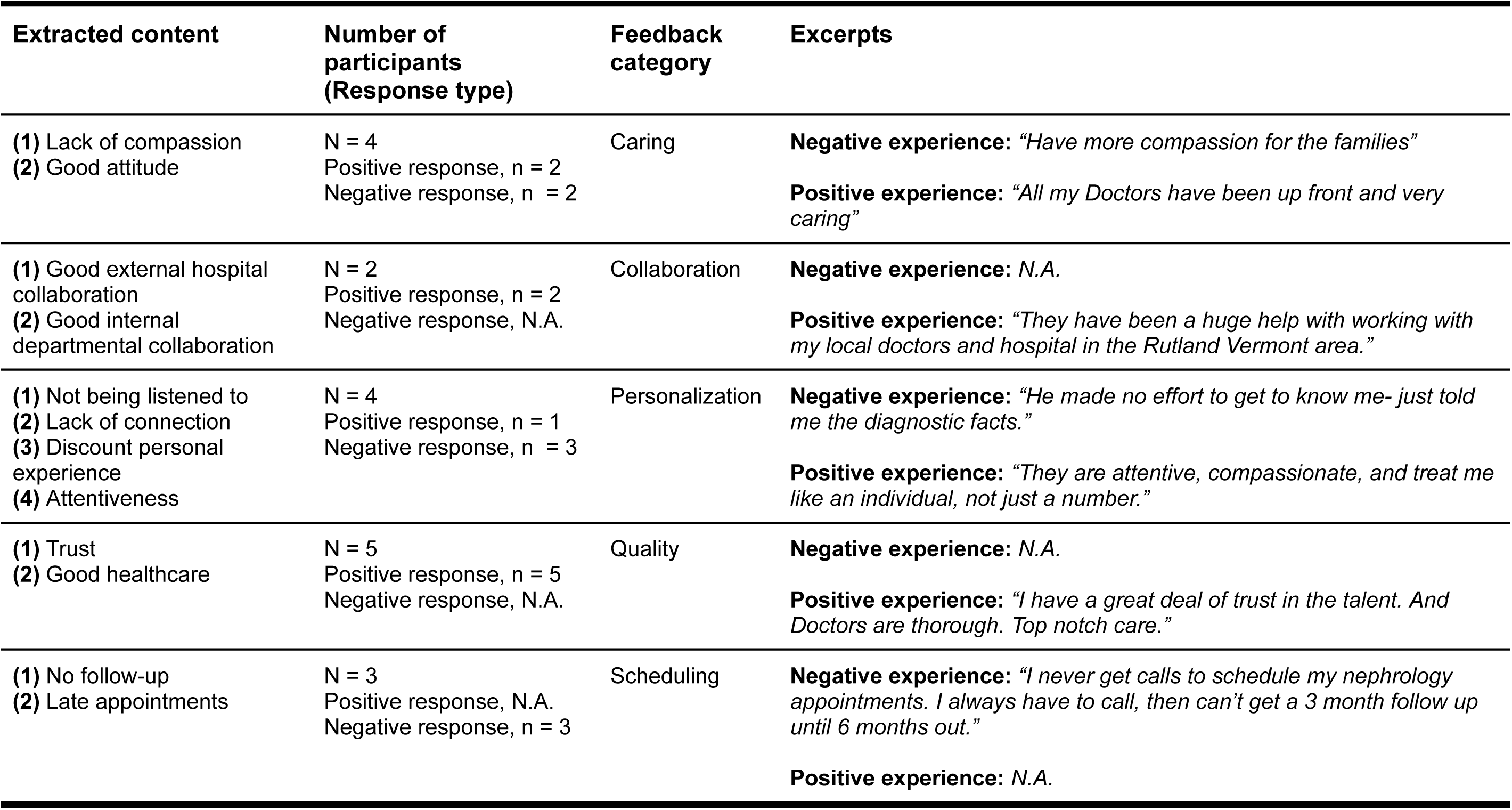
Content analysis based on participants’ response to the consideRATE open-ended question (N = 23).

**Table 6.** Measuring mean consideRATE scores across subgroups.

| **Subgroups** | **Mean considerATE score** | **t-value test statistic** | **df** | **p-value** |
| --- | --- | --- | --- | --- |
| Residence | Rural residing group: 3.61<br>Non-rural residing group: 3.68 | 0.916 | 96.47 | 0.362 |
| State | New Hampshire group: 3.70<br>Vermont group: 3.63 | 1.109 | 140.54 | 0.269 |
| Participant | Patient group: 3.62<br>Care partner group: 3.70 | 1.032 | 51.02 | 0.307 |
| Gender | Male group: 3.69<br>Female group: 3.61 | -1.145 | 150.02 | 0.254 |
| Reason for visit | Chronic kidney disease group: 3.69<br>Transplant or follow-up group: 3.60 | 1.288 | 147.89 | 0.199 |
| Distance traveled | Traveled less than 60 minute group: 3.57<br>Traveled at least 60 minute group: 3.72 | -1.77 | 93.77 | 0.080 |

Participants exclusively reported negative experiences in the categories Caring (n=2), Personalization (n=3), and Scheduling (n=3). We found issues like delayed appointments and lack of follow-up in scheduling frustrated patients and care partners. Likewise, patients and care partners reported a lack of compassion and effort by the care teams to get to know the patient individually as negative experiences. Conversely, participants only recorded positive experiences in the categories Collaboration (n=2) and Quality (n=5). Participants expressed strong trust in the quality of care at Dartmouth, as well as both Dartmouth’s interdepartmental collaboration and external collaboration with doctors in Northern New England. Positive experiences were also noted in the Caring (n=2) and Personalization (n=1) categories. We found that patients emphasized feeling valued when care teams attentively listened to participants and provided them with individualized, patient-centered care.

## Discussion

### Summary of results

This is the first real-world use of the **conside**RATE questions in a nephrology clinic among patients with kidney disease and their accompanying care partners. The **conside**RATE questions demonstrated reliability with strong internal consistency in addition to convergent and discriminant validity. For subgroup analysis, we found no significant difference in mean **conside**RATE scores across state residence, rurality status, gender, type of visit, responders,reason for visit, and distance traveled.

Our multivariable linear regression model revealed significant associations between healthcare experience measured by **conside**RATE and several demographic variables.

For instance, longer distance traveled and higher age were associated with higher **conside**RATE scores. People who were moderately religious, however, were likely to have lower consideRATE scores. Surprisingly, we found no significant association between healthcare experience measured by **conside**RATE and rurality measured with RUCA codes. For two **conside**RATE items, “What matters most” and “Surroundings”, we found that patients tend to rate (1) their healthcare providers’ attention to patients’ surroundings and (2) healthcare providers’ respect for them significantly lower compared to care partners. For the “Plans” **conside**RATE item, we found that participants who had a “high school degree or less” tended to rate healthcare providers’ communication about the next steps significantly lower compared to participants who had at least an undergraduate degree.

### Results in context

When we used the **conside**RATE questions in an outpatient nephrology context, our results were consistent with previous **conside**RATE psychometric studies from several other populations.^16,19^ The results in this real-world setting demonstrated strong psychometric properties, but they were less robust than in our online test with simulated patients.^16^ This makes sense, as the real-world context of a nephrology clinic introduces more variability and complexity than a controlled online simulation. For psychometric tests, our convergent validity, discriminant validity, and reliability tests in this nephrology clinical context were consistent with our findings in an outpatient cancer population, another real-world assessment.^19^

The association between rurality and patient experience has been inconsistent in previous studies.^22,24^ Our results contribute to this uncertainty in the literature by demonstrating that there is no significant (1) association between rurality and healthcare experience and (2) there is no significant difference in mean **conside**RATE scores between participants who live in a rural area compared to participants who live in a non-rural area. This lack of significant difference could be due to several factors. First, it is possible that healthcare services in this region, including access to care, have improved, reducing the disparities in healthcare experience that might have existed in the past. The expansion of telehealth services since the pandemic may have improved care at the hospitals and clinics that are left. Furthermore, Dartmouth-Hitchcock Medical Center, New Hampshire’s only Level 1 trauma center and a leading academic medical center in Northern New England, is situated in a rural area and provides high-quality care to rural patients that are nearby. Second, the majority of participants in this nephrology clinic reported “good” or “very good” when responding to **conside**RATE items. The homogeneity of responses (with a predominance of high scores) could imply that healthcare experiences across both rural and non-rural areas are perceived similarly, especially in areas where healthcare quality is generally high or where improvements in healthcare delivery have been consistent across regions. In such cases, rural-urban differences in healthcare experience may not be substantial enough to reveal significant differences in the **conside**RATE scores. Perhaps physical access and proximity to services is less important to patients than other aspects of their care.

### Implications

The **conside**RATE questions fill an important gap in the literature for assessing patient care experiences in chronic disease management, particularly in nephrology. CKD, for example, is a complex, lifelong condition that requires ongoing medical care, regular monitoring, and multidisciplinary support. In this context, it is critical to capture not only clinical outcomes but also the subjective experiences of patients, which can influence their engagement in treatment plans and overall sense of well-being. The **conside**RATE questions offer a unique approach by focusing on what matters most to patients in real-time healthcare interactions, making it particularly useful in chronic care settings where patient engagement and trust are essential.^34,35^

Our study has implications for multiple stakeholders in the care of patients with kidney disease. This measure allows patients to quickly and practically communicate elements of their experience to their clinicians, which can then inform management decisions with clinicians. With this measure, health systems can better track patient experience and implement policy changes to better facilitate quality patient care experiences.

Our **conside**RATE assessment revealed several key factors that may influence patient experience and could be addressed in nephrology clinics. For example, the significant difference observed in the “plans” item, where participants with a “high school diploma or less” rated their communication about next steps with healthcare providers lower than those with an undergraduate degree, perhaps demonstrates the importance of tailored communication strategies for patients and family members with serious illness, including those with CKD.^36^ This finding suggests that educational level may influence how effectively patients engage with their healthcare providers regarding their treatment plans. Furthermore, healthcare providers may need to adapt their communication style and ensure that patients with lower educational backgrounds receive clear and accessible explanations about their care plans. This includes ensuring that patient education materials are written at appropriate literacy levels, which can help enhance patient healthcare experience and satisfaction. Additionally, the lower ratings of the (1) “what matters most” and (2) “surrounding” **conside**RATE items suggest that patients may feel that their healthcare providers are not fully attuned to their personal environment, potentially impacting their overall experience of care. These results may suggest the need for healthcare providers to consider patients’ holistic needs, including emotional and environmental factors, in their care delivery.

### Limitations

As a single-center study, one significant limitation is that the population of Northern New England lacks the demographic diversity of other regions of the country. While the **conside**RATE questions may perform well in the population we studied, they may not be as readily understood by patients from other demographic groups, therefore limiting its generalizability.

Another limitation is that roughly 93 participants were excluded from the analysis due to interruptions in completing the measures in the waiting room of the nephrology clinic.

There is no clear reason to think that these excluded patients are different from those who were able to complete the measures before being called for their appointment. We note, however, that this limitation will be an important consideration as we design future studies with in-office recruitment.

## Conclusion

We demonstrated the first psychometric assessment of the **conside**RATE questions in a nephrology clinic. We found that patient experience did not differ by rurality in the population with CKD.

## Data Availability

All data produced in the present study are available upon reasonable request to the authors

